# Getting the Nod: Characterizing pediatric head motion in movie- and resting-state fMRI

**DOI:** 10.1101/2021.05.17.21257346

**Authors:** Simon Frew, Ahmad Samara, Hallee Shearer, Jeffrey Eilbott, Tamara Vanderwal

## Abstract

Head motion continues to be a major problem in fMRI research, particularly in developmental studies where an inverse relationship exists between head motion and age. Despite multifaceted and costly efforts to mitigate motion and motion-related signal artifact, few studies have characterized in-scanner head motion itself.

This study leverages a large public dataset (N=1388, age 5-21y, The Healthy Brain Network Biobank) to characterize pediatric head motion in space, frequency, and time. We focus on practical aspects of head motion that could impact future study design, including comparing motion across groups (low, medium, and high movers), across conditions (movie-watching and rest), and between males and females.

Analyses showed that in all conditions, high movers exhibited a different pattern of motion than low and medium movers that was dominated by x-rotation, and z- and y-translation. High motion spikes (>0.3mm) from all participants also showed this pitch-z-y pattern. Problematic head motion is thus composed of a single type of biomechanical motion, which we infer to be a nodding movement, providing a focused target for motion reduction strategies. A second type of motion was evident via spectral analysis of raw displacement data. This was observed in low and medium movers and was consistent with respiration rates. We consider this to be a baseline of motion best targeted in data preprocessing. Further, we found that males moved more than, but not differently from, females.

Significant cross-condition differences in head motion were found. Movies had lower mean motion, and especially in high movers, movie-watching reduced within-run linear increases in head motion (i.e., temporal drift). Finally, we used intersubject correlations of framewise displacement (FD-ISCs) to assess for stimulus-correlated motion trends. Subject motion was more correlated in movie than rest and stimulus-correlated stillness occurred more often than stimulus-correlated motion. Possible reasons and future implications of these findings are discussed.

## Introduction

Head motion remains a formidable challenge in fMRI research, particularly in developmental studies of functional connectivity [1,2]. Efforts to mitigate motion-related artifact are becoming both a subfield and an industry. Groups have tackled the problem from multiple angles: specialized behavioural training [3] and customized head molds have been designed to try to prevent motion [4], movies have been used to increase engagement, decreasing head motion and enabling longer functional runs [5–7], real-time motion monitoring systems allow researchers to index the amount of usable data collected as a scan proceeds [8], and techniques to mitigate head motion artifact are continuously evolving, including promising new approaches that leverage multi-echo sequences [9–11].

Some basic facts about pediatric head motion are well established: we know that children move more than adults, and that overall head motion in developmental samples almost always decreases with age. We also know that head motion, even in children, functions as a trait, just as it does in adults [12]. Beyond these basic attributes, and despite ongoing multifaceted efforts to decrease its effects, we know surprisingly little about pediatric head motion itself.

There are many developmental factors that suggest careful characterization of pediatric head motion is needed. In fMRI research, we usually take comfort in the fact that pediatric brain volume is roughly equivalent to adult brain volume by six years of age [13]. This makes using atlases and various software tools easier, but it also means that children’s heads are proportionally large relative to their bodies [14]. When lying supine in a scanner, a larger head necessarily causes differences in angles and weight distribution, for example, from the back of the head to the back of the shoulders. This means that a child’s head is more flexed when lying supine relative to adults [15]. The large head is supported on a neck that has relatively weak muscles and ligaments [16]. The biomechanical implications of these anatomical differences have been studied extensively in other disciplines such as helmet design and injury prevention [15,17], but the effect on head motion in the scanner is unclear.

Children also breathe differently than adults. They have smaller lung volumes, more flexible chest cavities, and larger tongues relative to the size of their oral cavity [18]. They are diaphragmatic breathers, meaning they use their abdomen and diaphragm during normal breathing to compensate for developmentally weaker intercostal muscles. Children also breathe faster than adults: normal respiratory rates for children ages 6-12 years are 14-22 breaths per minute [19]. Compared to adults who average 12-18 breaths per minute, this is a significant difference in movement frequency. When combined with a bigger head and more neck flexion at baseline, the effects of belly-breathing at a higher respiratory rate on head motion during scanning would theoretically be impactful.

Psychological factors are also relevant to pediatric head motion during tasks and movies, and especially during conditions in which adults typically mind wander [20,21]. Children generally do not talk about their own internal mental experiences as readily as adults or with the same level of “observership” or introspection [22]. In one study, children’s self-reports of mind-wandering were judged to be inaccurate, as their self-reports did not align with task performance or behavioural correlates [23]. When asked to try to have no thoughts for 20-25 seconds, 8-year-olds reported that they still had some thoughts, but most 5-year-olds reported having had no thoughts at all [24]. Children have also been shown to have different relationships between mental time travel and the use of cognitive resources compared to adults [25]. All enquiries about a child’s experience of spontaneous thoughts are confounded by developmental differences in language, and overall, the internal experience of children during tasks, movies and resting state is unclear. Theoretically, particularly during resting state, developmental differences in mind-wandering could have significant effects on head motion. For example, if it is true that children do not mind wander in an immersive or sustained manner in the absence of a task, they would likely become restless much more quickly.

Whether in the scanner or not, children move more than adults. Just as toddlers and young children thrive on repetition to drive their learning (e.g., hearing the same book over and over), the developing nervous system seems to require the constant input and output of physical activity. Studies of cortical thickness in infants ages 0-2 years show that somatomotor regions are already quite thin [26]. Even in a clinical MRI of an individual newborn infant, the somatomotor cortex stands out as being thin and looking mature relative to the rest of the cortex. Longitudinal mapping over an age range of 4-21 years shows that lower-order somatosensory and visual regions continue to mature ahead of higher-order association cortex as part of the “back-to-front” sweep of cortical thinning [27,28]. Functionally, the somatomotor network is also highly dynamic throughout childhood. From late childhood to early adulthood, nodes from the somatomotor network were most predictive of age [29,30]. As a child grows, and the nervous system is mapped and remapped onto a changing physical structure, the somatomotor network exhibits dynamic structural and functional changes. The effect of these changes on movement patterns in general, and on head motion in the scanner in particular, is unclear.

In sum, there are anatomical, physiological, psychological and neural reasons that head motion in children is high and inversely related with age, yet we do not know how these features affect head motion during scanning. Here, we try to fill this knowledge gap by characterizing multiple aspects of head motion in children and youth using a large publicly available developmental dataset (The Healthy Brain Network Biobank) [31]. We focus on practical aspects that might have implications for study design, or that might inform efforts to prevent motion or to mitigate motion-related artifact. We characterize head motion in space, frequency and time. Comparisons are made across conditions (movie-watching and rest), and across movement cohorts (high-, medium- and low-movers). We leverage a large sample size to test for sex-based differences in head motion, and finally, we assess for stimulus correlated motion trends.

## Methods

### Sample

Data from the Healthy Brain Network biobank (HBN) were used for all analyses [31]. This public dataset contains imaging data and phenotypic measures for transdiagnostic research in children and adolescents (age 5-21y) from the greater New York area. All participants were seeking psychiatric help. The Chesapeake Institutional Review Board approved all study procedures. Written consent was obtained from participants aged 18 years and older, while parental consent and written assent were obtained from all other participants.

Here, data from 2041 subjects were initially accessed and N=1388 subjects were retained after excluding those with incomplete demographic data, missing volumes, or absence of one of the three functional runs of interest (491 females, 5-21y, mean age 11.0 ± 3.4y, HBN-1388). The Child Behaviour Checklist (CBCL) was available for 865 participants and was used to reflect broad levels of psychopathology and behavioural problems (n=865, 317 females, ages 5-21y, mean age 10.8 ± 3.15y, HBN-865). See Fig 1 for sample demographics and distributions.

**Fig 1.**
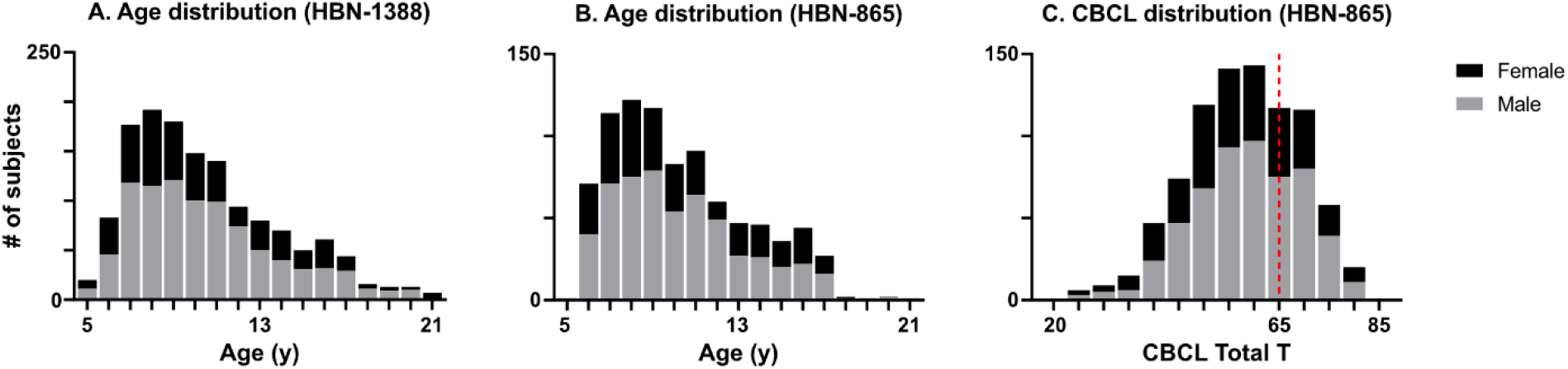
Sample demographics. **A**. Age and sex distribution of HBN-1388 (491 females), mean age 11.0 ± 3.4 years. **B**. Age and sex distribution of HBN-865 (317 females), mean age 10.8 ± 3.15 years. **C**. Child Behaviour Checklist (CBCL) Total T-scores, a questionnaire to assess general psychopathology and behavioural problems. Higher scores indicate more problems; the right skew here suggests psychiatric enrichment. The red line shows the general threshold for clinical concern, which is 65.

### MRI acquisition

MRI data were collected at either Rutgers University using a Siemens 3-Telsa Tim Trio or at the Cornell Brain Imaging Center using a Siemens 3-Tesla Prisma. Echo-planar imaging sequences for all functional runs used TR=800ms, TE=30ms, FA=31°, slice thickness=2.4mm, FOV=204mm, a multiband factor of 6, and voxel size=2.4×2.4×2.4mm.

### Scanning sessions

The HBN scan sessions were 64.7-minutes long. First, an initial localizer, one or more T1-weighted anatomicals (HCP T1 with *Inscapes* being shown and/or ABCD T1), and an EPI Field Map were run [7]. Next, three functional runs were interspersed with predictive eye estimation regression (PEER) calibration runs (Rest1, PEER1, Rest2, PEER2, Movie-F [*Despicable Me*]). After Movie-F, diffusion kurtosis imaging (DKI), DKI-field mapping, and additional T1/T2 anatomical runs were completed before a final functional PEER session and second movie-watching run (*The Present*) [31–33].

Three functional runs were used in this study: two 5.1-minute runs of eyes-open rest during which a fixation cross was displayed (Rest1, Rest2, 375 volumes each) and one 10-minute run during which a clip with audio from the movie *Despicable Me* was shown (Movie-F, 750 volumes) [32]. The ten-minute scene from the movie was chosen to be emotionally charged. It features a poignant attachment-based scene in which orphaned children convince their begrudging caregiver to read them a bedtime story. It ends with a verbal argument and moral conflict for the main character, and finishes with a comical scene featuring nonverbal minions and a photocopy machine. Data from Movie-F was truncated to match the number of volumes in Rest1 and Rest2, creating Movie-H (Movie-F = Movie-Full, Movie-H = Movie-Half). The first and last 10 volumes of all conditions were excluded, leaving either 355 or 730 volumes for analysis.

### Preprocessing

The MCFLIRT motion correction tool (FSL 6.0) was applied with default settings, encoding motion as displacement from the middle volume, yielding a six-dimensional time series (translation: x, y, z, rotation: pitch, roll, yaw) per subject and per run [34].

### Analyses

#### Data availability

Data are from the Healthy Brain Network Biobank and are available at http://fcon_1000.projects.nitrc.org/indi/scmi_healthy_brain_network/ and are subject to a data usage agreement. Specific to this study, subject IDs and code are available at https://github.com/tvanderwal/head_motion_2021. All computations and analyses were carried out using MATLAB R2019b and GraphPad Prism version 9.1.0 unless otherwise specified.

#### Motion parameters

Framewise displacement (FD) was calculated per volume by backward difference as in Power et al. and rotations were converted from radians assuming a 50mm radius sphere [1].

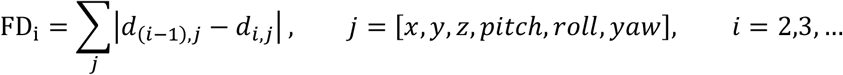

Each subject was classified as a low, medium, or high mover for each condition based on mean FD as follows: Low: < 0.15mm, Medium: 0.15 - 0.3mm, High: > 0.3mm. These thresholds were selected to reflect common standards in the field (e.g., Fair et al. [35]).

To assess motion within each of the six rigid-body axes, the FD formula was applied without the final summation step, yielding a six-dimensional time series of relative differences.

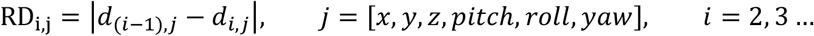

To determine the percent composition of FD with respect to rigid-body axes (i.e., the contribution of motion from each axis to the mean FD), the value for each axis was divided by FD per volume. Since FD is the sum of all axes, this converts the value to a proportion and yields a six-dimensional time series of FD percent composition.

#### Motion spikes

A motion spike was defined as a volume with FD greater than 0.3 mm. The FD from motion spike volumes was averaged to generate mean spike FD per subject for each run. To assess the type of motion occurring during motion spikes, the rigid-body composition of motion spikes was also extracted and averaged per subject for each run as above.

#### Frequency analysis

To characterize the frequency of movement in these data, the absolute displacement from each axis was transformed into power spectral density using the *pmtm* function in MATLAB with a time-half bandwidth product of 8 and 512 points, log transformed, and z-scored. These procedures closely followed Fair et al. [35].

#### Motion drift

To visually assess within-run and within-scan motion trends over time, mean FD by volume was calculated within each motion group for each condition. To enable statistical comparison of motion drift across conditions, a linear regression of FD was performed and the slope of each condition per subject was retained and compared. A positive slope indicates a linear increase in FD during a run.

#### Stimulus-correlated motion trends

A sliding window approach was used to assess for intersubject correlations in FD time series. FD time series were subset into 15 volume (12 second) windows and correlated across subjects for each condition. Resulting Pearson’s correlation coefficients (r values) were Fisher Z-transformed and averaged to generate a representative value of FD-correlation per window. The choice of 12-second windows was loosely based on the shortest scene length we thought might be identifiable in the movie during subsequent reverse annotation. This length of window was not meant to identify tight, stimulus correlated movements (e.g., jolting in response to a jump-scare in a horror movie), but rather to identify general trends in motion that were shared across subjects during epochs of the movie.

## Results

### Basic attributes of mean FD

#### Across conditions

Statistically significant differences in mean FD were found across all condition pairings except for Rest1 to Movie-F. When comparing conditions of the same duration, mean FD during Movie-H was lower than both Rest1 and Rest2, and even the ten-minute long Movie-F had lower mean FD than Rest2 (one-way repeated measures ANOVA with Geisser-Greenhouse correction to account for non-sphericity (F(1.896, 4161)=52.35, p<0.0001) with follow-up two-tailed t-tests Bonferroni corrected for six comparisons (p<0.0083)). *<u>With age:</u>* As expected, mean FD was negatively correlated with age in all conditions (Rest1: −0.31, Rest2: −0.34, Movie-H: −0.25, Movie-F: −0.27, all p<0.0001). *<u>With CBCL:</u>* No significant correlations were found between mean FD and CBCL in any condition (r (p), Rest1: 0.055 (0.11), Rest2: 0.052 (0.13), Movie-H: 0.02 (0.48), Movie-F: 0.018 (0.60)). *<u>Motion groups:</u>* Three motion groups were defined for each condition using mean FD cut-offs of 0.15mm and 0.3mm. Significant differences in age were found between all motion groups per condition, and between CBCL for low-high and medium-high comparisons in Rest1 and Rest2 (For each condition and measure pairing, eight total: Kruskal-Wallis test with follow-up Dunn’s tests Bonferroni corrected for three comparisons, p<0.0166). The Kruskal-Wallis (comparison of medians) and Dunn’s tests (comparison of rank sums) were chosen as a non-parametric alternative for non-gaussian data distributions. Overall, there were more high movers than low movers, and high movers were younger. CBCL was significantly different for the low-motion group in both Rest1 and Rest2, but not in Movie-H or Movie-F (though the trend was directionally the same). More than half of participants (n=774, or 56%) were in the same motion group for all conditions. See Fig 2 for results.

**Fig 2.**
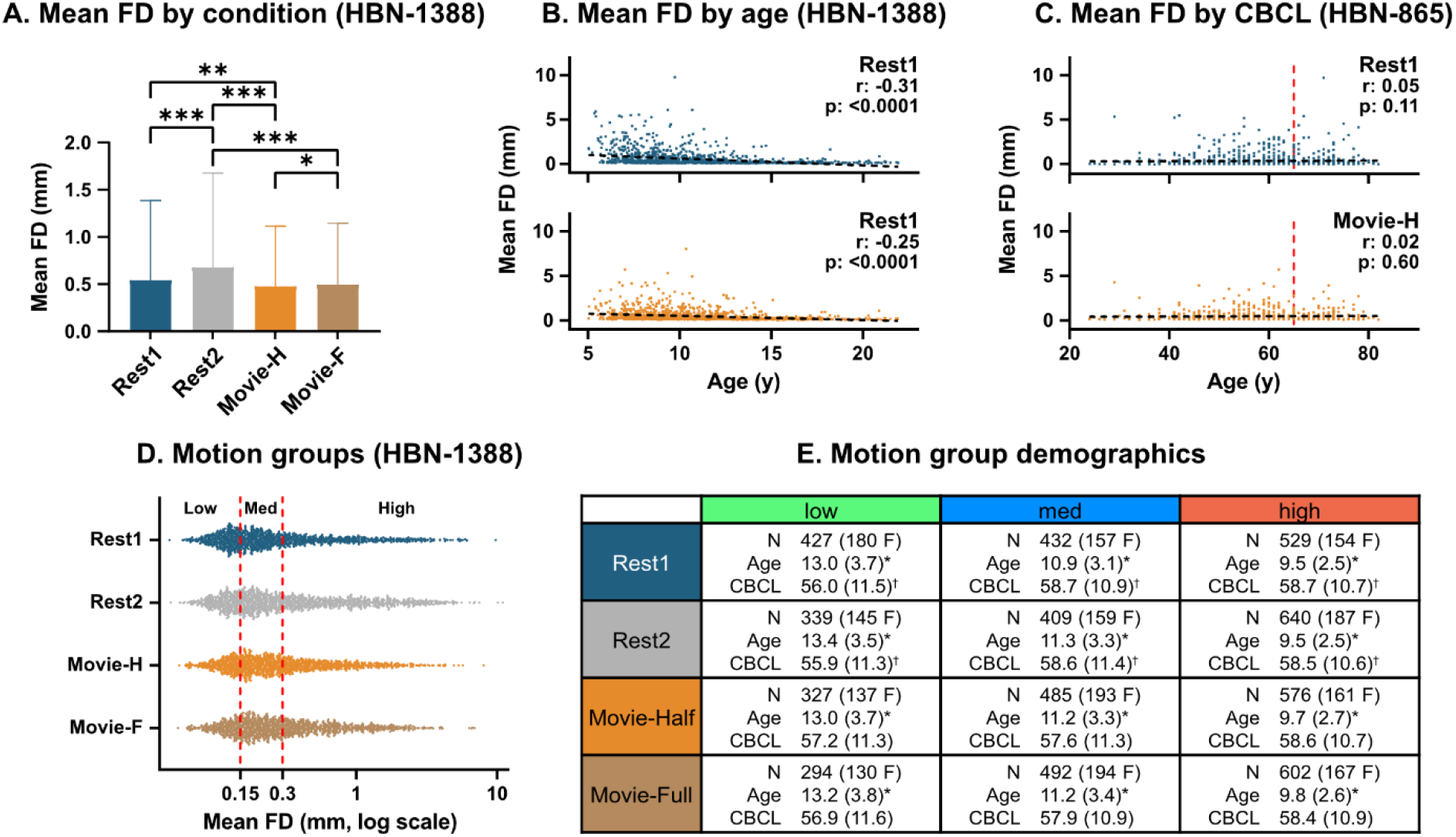
Head motion summary statistics. **A**. Mean FD by condition: significant differences between all conditions except for Rest1 and Movie-F (*** = p<0.0001, ** = p<0.01, * = p<0.05). **B**. Mean FD by age: significant negative correlations in all conditions. **C**. Mean FD by CBCL: no significant correlations in any condition (cut-off for clinical concern is 65, red line). **D**. Motion group distribution by condition. **E**. Motion group demographics by condition: significant differences were found for age across groups (asterisk) and for CBCL across low-high and medium-high groups (dagger).

### Motion composition by axis

To visualize trends in FD composition from low to high movers in a continuous manner, percent mean FD contribution from each of the six axes was calculated. Subjects were sorted by mean FD, and percent mean contribution values were smoothed using a 200-subject moving average. Fig 3A shows the smoothed FD composition versus mean FD in Rest1. In low movers, motion was primarily composed of y > z > pitch axes. High movers showed a different pattern that was dominated by pitch with lower translation along the y- and z-axes (i.e., most of the motion occurred in the same three axes, but the pattern was flipped for high movers).

**Fig 3.**
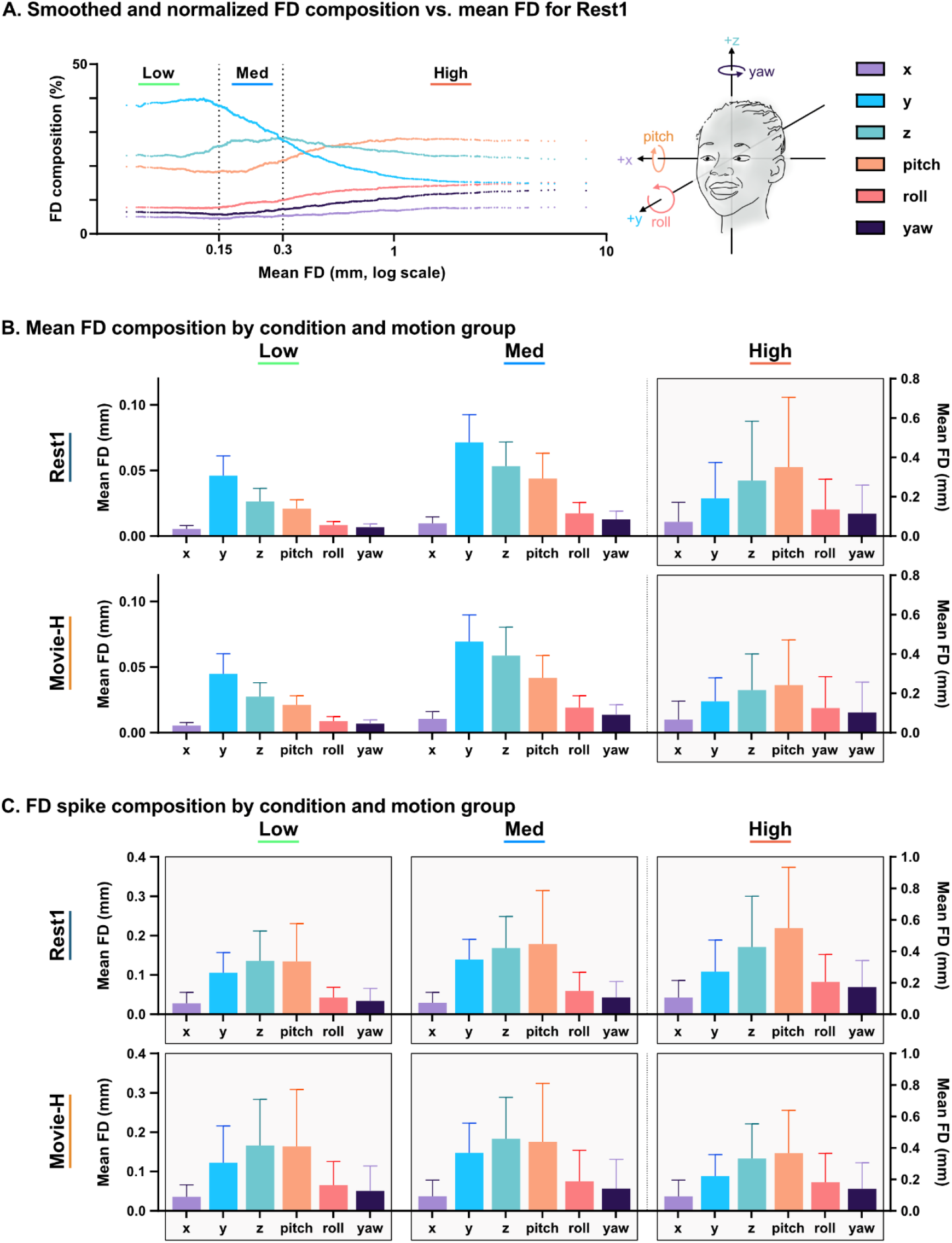
Head motion composition. **A**. Mean FD percent composition: Contribution of rigid-body axes to mean FD shifts as motion increases (y > z > pitch to pitch > z > y). **B**. Mean FD composition: Significant differences were found between low-high and medium-high motion groups both within and between all axes within each motion group for both conditions. **C**. FD spike composition: Significant differences as above except that z-translation and pitch were similar within-group for low-medium comparisons. The high-movement pattern in both high movers and spikes is highlighted by grey boxes in 3B and 3C.

Quantitative analysis showed statistically significant differences in mean FD for both Rest1 and Movie-H between all axes within each motion group, and between low-high and medium-high groups within each axis. Overall, high movers had a different motion signature than medium and low movers regardless of condition (Fig 3B). Statistical testing was conducted for each condition as follows: three repeated measures ANOVAs with Geisser-Greenhouse correction between axes for each group and six ordinary ANOVAs between groups for each axis, all p<0.0001 with follow-up two-tailed t-tests (Bonferroni corrected for 63 comparisons).

To quantify motion composition of high-motion spikes (rather than mean FD), we identified all volumes with mean FD > 0.3mm, computed FD contribution from each axis for each spike, and then averaged to generate mean spike FD composition per subject. This was then averaged at the group level to produce a per-axis summary measure of high-motion volumes for each condition. Within each motion group, statistically significant differences in spike composition were found across all six axes, with the exception that z-translation and pitch were similar in low and medium groups. Within each axis, differences were also found between low-high and medium-high movers regardless of condition. In sum, though high movers and low movers had different movement patterns when looking at mean FD, all high-movement spikes—from any movement group or in any condition—showed the same movement composition as high movers (Fig 3C). Statistical testing was completed as mentioned previously for mean FD composition with nine ANOVAs and follow-up Bonferroni corrected two-tailed t-tests.

### Frequency analysis of raw displacement data

Differences between low, medium, and high movers were also evident when the frequency of motion was analysed. Low and medium movers demonstrated greater spectral power in the y- and z-axes in the 0.2-0.4 Hz range (12-24 events per minute), whereas this pattern was not observable in high movers. These findings are shown for resting state (Fig 4) but were highly similar across all conditions, and also replicate findings by Fair et al. [35].

**Fig 4.**
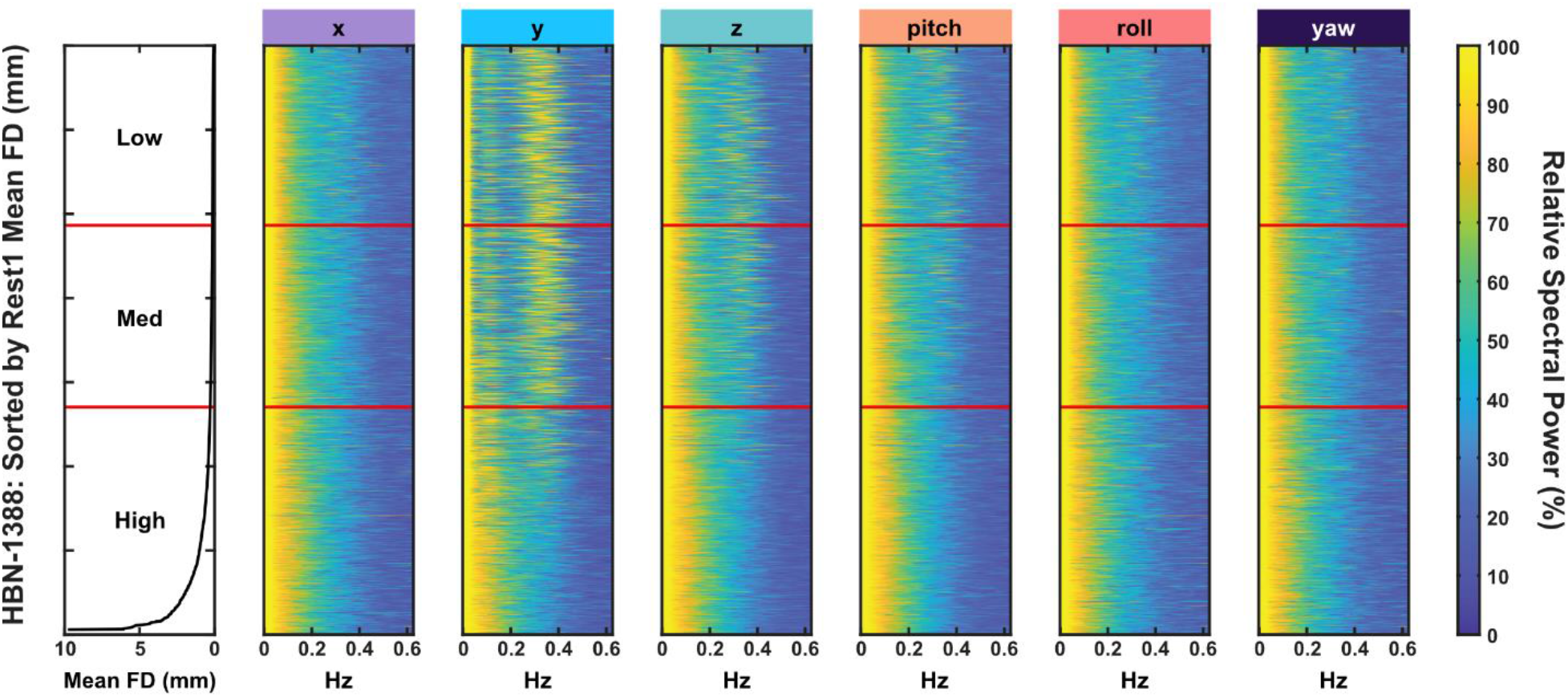
Frequency analysis of raw head motion. Greater spectral power is observed in the y- and z-axis in low and medium movers in the 0.2-0.4 Hz range (12-24 events per minute). This is not evident in high movers. Data shown is from Rest1, but highly similar results are found across all conditions and replicate findings by Fair et al. [35].

### Temporal drift in mean FD

Mean FD by volume was plotted for each functional run to qualitatively assess motion trends over time. Mean absolute deviation of volume-wise FD (MAD) was chosen as a lower-magnitude alternative to standard deviation with which to show variability 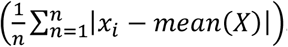. A positive linear drift in mean FD is apparent in high movers during Rest1 and Rest2, but not in Movie-H or Movie-F (Fig 5A). To statistically test for cross-condition differences in drift, a best-fit line (i.e., slope) was computed for each subject in each condition from their mean FD by time graph. Statistical tests conducted using these slopes showed significant differences in slope across conditions in the medium and high movers, and in general, motion drift was greater during five minutes of rest relative to five or ten minutes of movie-watching (three Kruskal-Wallis tests, one per group: Low: H(3)=18.82, p=0.0003, Med: H(3)=28.48, p<0.0001, High: H(3)=126.4, p<0.0001, with follow-up Dunn’s tests Bonferroni corrected for six comparisons, p<0.0083, see Fig 5B).

**Fig 5.**
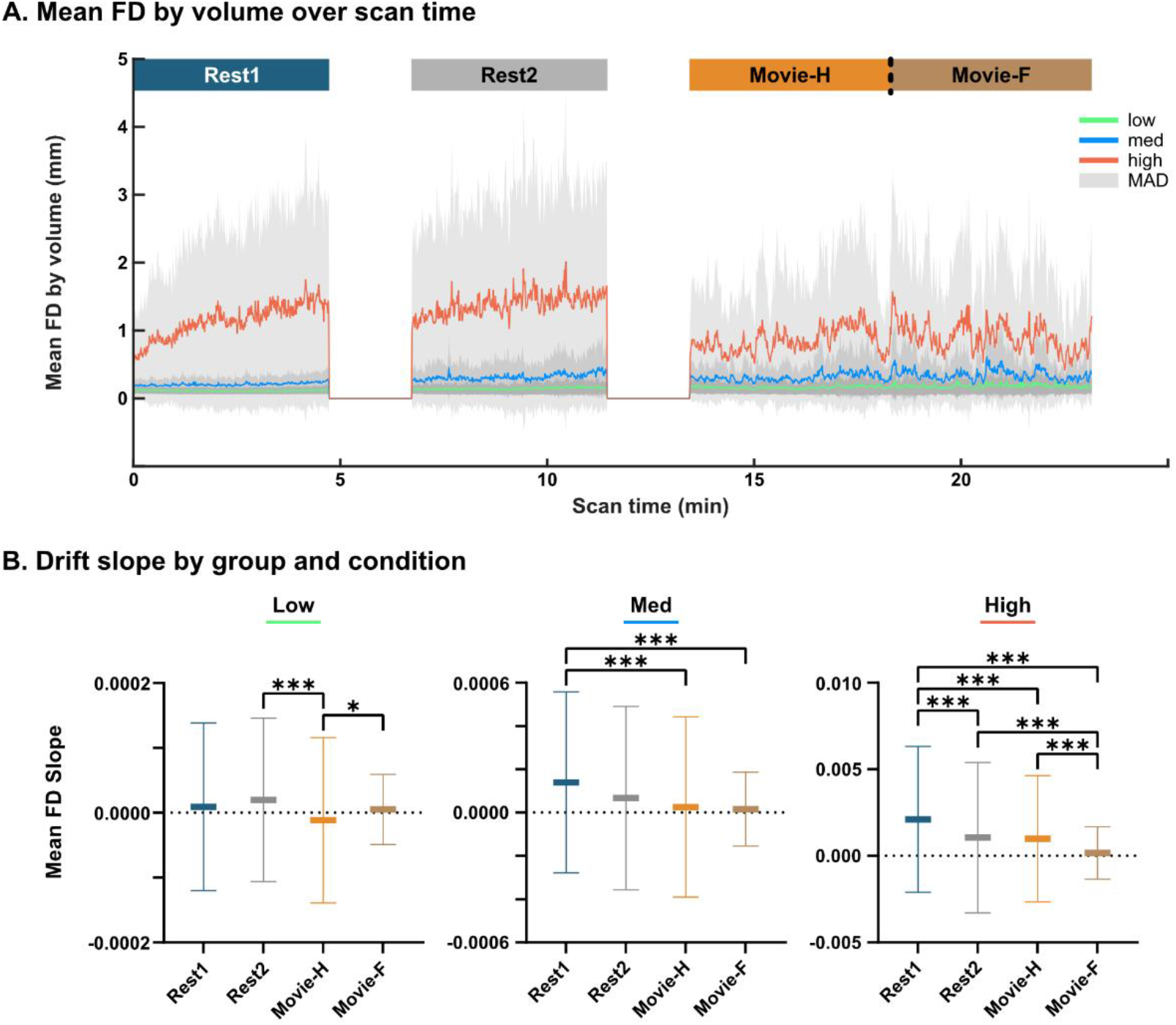
Temporal drift in mean FD. **A**. Mean FD by volume over scan time: Mean FD by volume and mean absolute deviation (MAD, grey) was calculated for each motion group and condition. Data show a linear temporal drift in high movers in Rest1 and Rest2 but not in Movie-H or Movie-F. **B**. Drift slope comparison: High movers had the largest differences in within-run temporal drift (i.e., slope) between conditions with all comparisons being statistically significant except for Rest2 to Movie-H (*** = p < 0.001, * = p < 0.0083). The scale of slopes was more than an order of magnitude greater in high movers relative to low and medium movers.

### Sex-based differences in FD

The large sample size enabled us to assess for sex-based differences in head motion. When split by males and females, there was no difference in mean age (unpaired two-tailed t-test: t(1386)=0.109, p=0.913) but males had significantly greater CBCL scores relative to females (unpaired two-tailed t-test: t(863)=2.413, p=0.016) (Fig 6A). Females had significantly lower mean FD in all conditions (Kruskal-Wallis test: H(7)=117.4, p<0.0001, with Dunn’s tests Bonferroni corrected for four comparisons, p<=0.0002) (Fig 6B). ANCOVAs were performed to assess for an age-by-sex interaction, but no interaction was found (Rest1: F(1, 1384)=0.16, ns, Movie-H: F(1, 1384)=0.62, ns). To look for sex differences in motion composition (both in mean FD and high-motion spikes) percent axial composition of mean FD and mean spike FD was calculated for females and males for Rest1 and Movie-H (Fig 6C). Qualitatively, motion composition appears similar across males and females, and Dunn’s tests across sex within each condition showed no significant differences between axes (Four sets of six Dunn’s test for each condition and motion-type combination, Bonferroni corrected for six comparisons). In sum, males moved more than, but not differently from, females in this sample.

**Fig 6.**
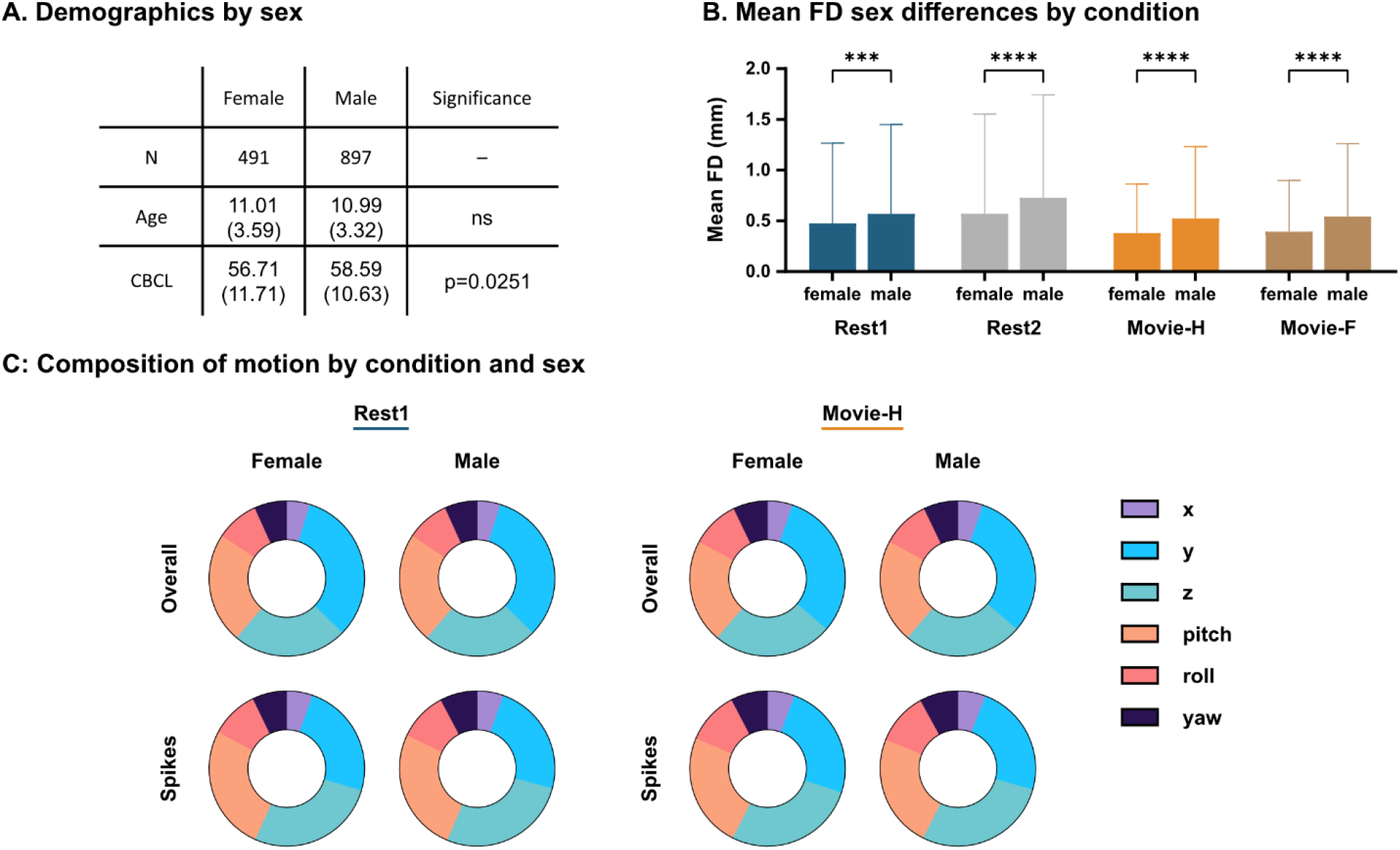
Sex differences in FD. **A**. Demographics by sex: Sample is heavily male-weighted, age is not different, and males have greater CBCL scores, which indicates greater psychopathology and/or behavioural problems. **B**. Mean FD sex differences by condition: Across all conditions, head motion was lower for females than males. (**** = p<0.0001, *** = p=0.0002) **C**. Motion composition by condition and sex: Percent composition for both mean FD and FD spikes was not significantly different across sex in either condition, indicating that males moved more than, but not differently from, females.

### Stimulus correlated motion

To assess stimulus correlated motion trends during the movie, we computed intersubject correlations of FD (FD-ISCs) using a sliding window approach. Mean Fisher z-transformed r-values (z’) during movie-watching were low and ranged from 0.025-0.003. Despite the low absolute magnitude, these were significantly greater than during Rest1 or Rest2, which ranged from 0.002-0.007. Differences in FD-ISCs were statistically significant across all pairings (Kruskal-Wallis test: H(2)=661.8, p<0.0001, follow-up Dunn’s tests Bonferroni corrected for three comparisons, all p<0.0001)) (Fig 7A). To relate FD-ISCs to mean FD, we then computed mean FD using the same window length. The top ten FD-ISC peaks during Movie-H were marked, and the windowed mean FD during that epoch was classified as being above or below the mean FD for the whole run. Eight of the ten more synchronized epochs occurred when head motion was below the mean, suggesting times of stimulus correlated stillness. To see if particular types of scenes were occurring during the correlated stillness or correlated motion windows, we looked at what was happening in the movie during the ten more synchronized epochs (i.e., reverse-inference) (Fig 7B). These epochs overlapped and clustered onto focused, highly social, dialogue-heavy scenes featuring close-up camera views and low local motion, such as the bedtime story scene. The two epochs with FD above the mean occurred during an argumentative scene in which the main character experiences a moral dilemma and there is a sense of discomfort or threat. Overall, a clear negative correlation was found between FD-ISC and FD (r=−0.49, p<0.0001) (Fig 7C).

**Fig 7.**
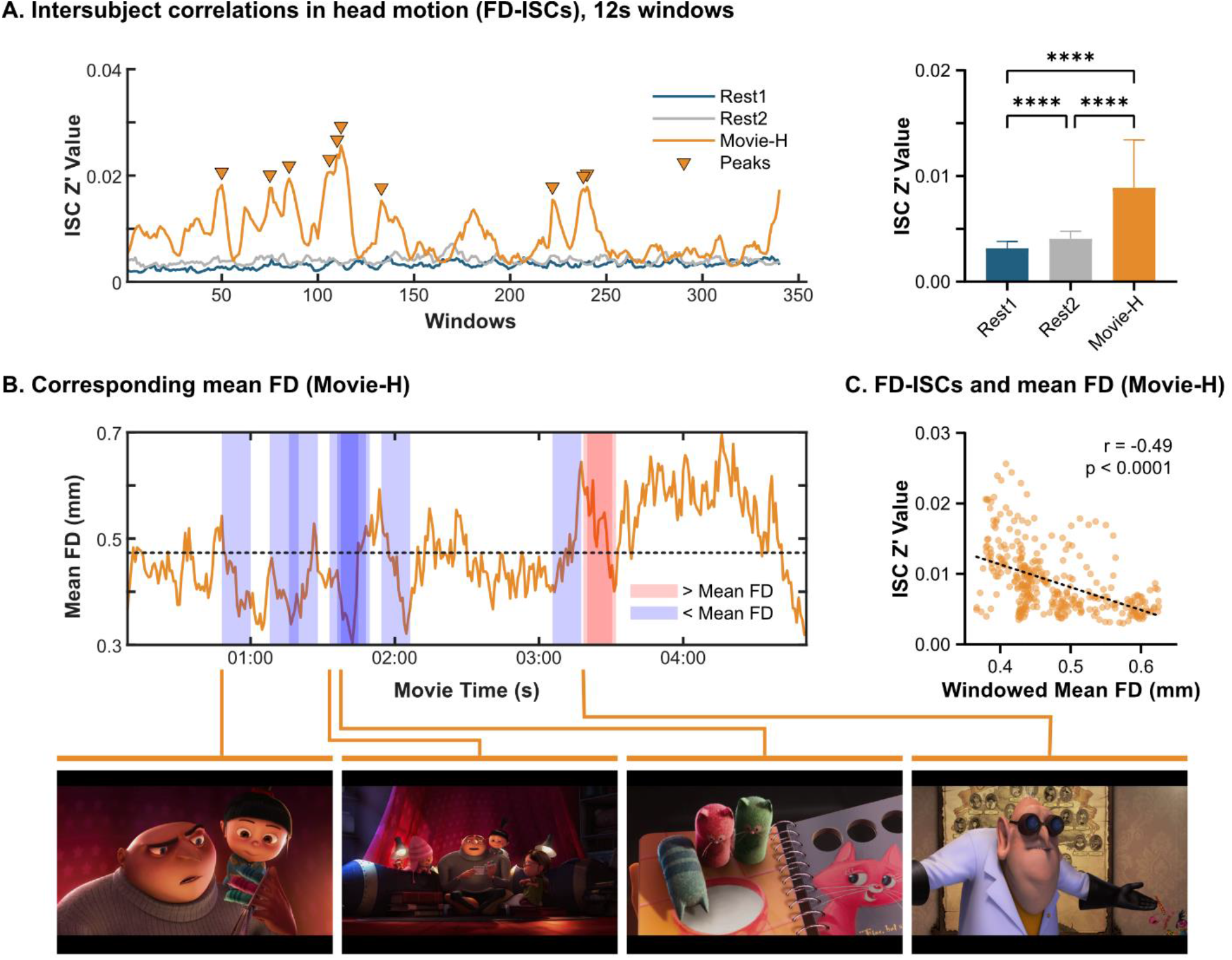
Stimulus correlated motion. **A**. Intersubject correlations in head motion (FD-ISCs): Movie-H mean FD-ISC is significantly higher than Rest1 and Rest2 (**** = p<0.0001). **B**. Corresponding mean FD time series: The top ten FD-ISC peaks were identified, and the corresponding windows were overlaid on the volume-wise mean FD time series for Movie-H. Red windows contain mean FD that is above the overall mean, blue shows windows in which FD was below the mean. Representative frames from these epochs are shown - epochs clustered into highly social, close-up, dialogue-heavy scenes. **C**. FD-ISCs and mean FD: Windowed mean FD was negatively correlated with FD-ISC values, indicating that stimulus-correlated motion trends during movie-watching were mainly driven by lower head motion or stillness.

## Discussion

Head motion during fMRI scanning causes artifactual signal changes that are an inherent, age-related confound in developmental analyses, especially using functional connectivity (FC) data. Here, we characterized head motion in a large developmental database (N=1388), aiming to better understand practical aspects of in-scanner head motion in space, frequency and time. We focus on cross-condition differences in framewise displacement (FD), and also comparisons across low-, medium-, and high-movement cohorts.

### Problematic head motion has a consistent signature

The results of multiple analyses of head motion composition (i.e., the contribution to mean framewise displacement from the six rigid-body axes) were remarkably clear, showing that across conditions and groups, higher in-scanner movement is dominated by rotation around the x-axis (pitch) and translation along the z- and y-axes. This was true of mean FD in the high-movement cohort, and of all high-movement spikes regardless of group or condition. We interpret this pitch-z-y pattern of movement as resulting from a simple nodding motion (flexion and extension of the cervical spine) and suggest that this provides a simplified target for interventions to reduce in-scanner head motion.

Why might this particular nodding movement be happening in the scanner? Because of their large head-to-body ratio, children are already in a degree of neck flexion when lying supine in the magnet. The nodding motion might be an effort to regain a neutral neck position, from which they naturally fall back into flexion. It would be interesting to see if using a foam pad under the body of participants that have higher head-to-body size ratios to create a neutral cervical spine during scanning would decrease head motion. Another possible explanation relates to psychological processes during scanning. We hypothesize that children may be increasing neck flexion to try to look outside the bore of the magnet or down at their bodies, possibly to make sense of their environment, out of boredom, curiosity, or some natural instinct that occurs when one is lying inside a tube. If this is the case, a curtain at the end of the bore might be helpful for some children. Available binocular display goggles that are worn on the face and restrict field of view to the stimuli alone may also be of benefit.

We considered whether the nodding movement might occur in the context of the type of diaphragmatic breathing that children do, but spectral analysis of head motion suggested that this is not the case. We observed low-frequency movements (0.2-0.4Hz) in the y- and z-axes in low and medium movers but not in high-motion subjects. These findings replicate previous work by Fair et al. who also interpreted this pattern as being consistent with respiratory movement. The fact that the respiratory pattern is not seen in high movers most likely means that it is obfuscated by different and greater motion patterns, specifically the pitch-z-y movement of interest. In our view, low-frequency breathing motion comprises a physiological minimum for in-scanner head motion that is a non-modifiable variable best targeted during data preprocessing.

The specific flexion/extension movement might also be amenable to physical restriction via more targeted devices. For example, it is possible that a soft neck brace to prevent flexion might be as effective as a full headcase. Overall, in this large, psychiatrically enriched developmental sample, “bad” head motion presented as a focal, specific type of movement, rather than as a global, idiosyncratic issue involving the whole range of biomechanically possible movements. Though the movement itself may be consistent across subjects and conditions, it is likely that (as in all things pediatric), developmentally appropriate and individualized approaches to decreasing this movement will end up being the most effective overall.

### Effect of movie-watching on head motion

Movies showed specific advantages in head motion. As previously shown, clear cross-condition differences in mean head motion were shown, and particularly in the high-movement cohort, movies made a significant difference in decreasing head motion [6,7]. New findings shown here are that movies prevent a linear drift in mean head motion over time, an advantage that persists even when the scan duration of the movie is doubled (ten minutes) relative to rest (five minutes). Again, this advantage was most prominent for the high movers, for whom linear drift within even a 5-minute resting state run was significant.

We also assessed stimulus correlated motion using an intersubject approach. Mean intersubject correlations of framewise displacement (FD-ISCs) were low overall, but were statistically greater during movies relative to rest. A clear negative correlation was shown between FD and FD-ISCs. Further, of the ten most correlated epochs, eight of them occurred when FD was below the mean for the whole movie. This suggests that, for much of the time, movies evoke stimulus correlated stillness. When we looked back at the movie to see what types of scenes were happening during the stillness, the epochs clustered around one of the more intimate, dialogue-heavy scenes with close camera angles, and low local motion (the storybook scene). The two epochs with FD above the mean occurred during an argumentative, tense scene. It may be possible to leverage these findings to select or create movies that evoke stimulus correlated stillness for longer periods of time. Additionally, future work is needed to more fully understand both stimulus correlated stillness and stimulus correlated motion. Stimulus correlated motion may be especially problematic as a source of systematic artifact, and for example, it may be helpful to apply a movie-based version of motion censoring in which epochs of data with correlated motion are removed. First, though, more exact methods than the intersubject sliding window approach used here are probably needed to better understand stimulus correlated motion.

### Other factors

In this sample, head motion was not correlated with scores from the Childhood Behavior Checklist (CBCL) which we used as a measure of general psychopathology and behavioural problems. This is in contrast to the other large-scale study of pediatric head motion (1134 scans, ages 10-16 years), where patients with defined disorders such as autism spectrum disorder and attention-deficit hyperactivity disorder (ADHD) and at-risk participants had significantly greater FD than controls [8]. Even in that study, though, participant demographics explained only 12% of the variance in head motion. Other disorder-specific studies have found no relationship between diagnosis and head motion. For example, no difference in head motion was found between youth and adults with ADHD and age-matched controls (Epstein 2007). Overall, the relationship between head motion and psychiatric disorders is not clear cut and generalizable across studies and symptom measures, and in this study, using a generalized “dimensional” measure of behavioural problems (CBCL), we did not find a relationship with head motion. We would agree with Dosenbach et al. [8], who concluded that head motion is driven more by individual factors.

In this sample, females moved less than males across all conditions. This finding is consistent with other large samples [8], but is in contrast to smaller studies that found no difference in head motion between males and females [5,12]. Also, no sex-by-age interaction was found with regard to head motion. This is not that surprising, as females are generally developmentally ahead of males during childhood and adolescence [36–38], so chronological age is likely a poor index in this case. Though females in this sample moved less than males, the motion they exhibited was compositionally the same as males. Just as the relationship between age and head motion is an inherent confound for developmental studies, this very clear female/male finding may represent an important confound for which future studies on sex-based differences will need to account.

### Limitations

There are multiple limitations to this study. First, all analyses were conducted using standard measures of mean framewise displacement generated via FSL, and we did not run analyses using other complementary measures such as DVARS. Despite a focus on head movement and developmental differences, we did not use subject-specific head circumference when computing measures of rotation, which might be worth doing in the future. Further, it would be interesting to probe the relationship between head circumference and mean FD, particularly in participants with larger head circumferences. The interpretation of the main finding, that most problematic head motion is composed of a signature pitch-z-y movement consistent with a nodding motion is, at this point, inferential. Future studies could look at framewise displacement from intentional nodding movements to test this interpretation. We also do not make any child/adult comparisons, which might also be warranted in future work. There are major limitations to the stimulus correlated motion analysis. In particular, because movies contain so much dynamic complexity, reverse annotations such as performed here are largely inferential endeavors. Due to the small sample of epochs used, our observations about what types of scenes engender stillness may not be generalizable, and further prospective study is needed.

## Conclusions

1. High head motion demonstrated a consistent pattern that was primarily composed of rotation around the x-axis, and translation along the y- and z-axes. This pitch-y-z combination of motion is consistent with a nodding movement, and it was the signature of mean framewise displacement in the high movement cohort and for all high motion spikes regardless of motion group, condition, or sex. This single biomechanical movement provides a focused target for motion reduction strategies.
2. Low-frequency movements (0.2-0.4Hz) consistent with breathing were observed in low and medium movers as translation along the y- and z-axes. We consider this to be a non-preventable source of movement that is best targeted in post-processing.
3. Females moved less than males during all conditions, and there was no sex-by-age interaction. This factor should be assessed in future FC studies, particularly of sex differences.
4. Movies had a significant effect on head motion, and mean FD for the movie was significantly lower than for rest in the sample as a whole. This advantage stemmed in large part from within-run linear increases in head motion that occurred during rest. This temporal drift affects higher movers preferentially, and was observed even in the very first 5-minute run of rest. Conversely, temporal drift in FD during the movie was remarkably low, even when the movie was twice as long as rest, and occurred later in the scanning session.
5. The movie resulted in greater intersubject similarity in FD compared to rest. These intersubject correlations in FD (FD-ISCs) were negatively correlated with FD, and the most correlated epochs had FD that was below the mean. We conclude that movies evoke stimulus correlated stillness during key scenes, which may contribute to the lack of a temporal drift in FD, and to lower FD overall.

## Data Availability

Data are from the Healthy Brain Network Biobank and are available at http://fcon_1000.projects.nitrc.org/indi/scmi_healthy_brain_network/ and are subject to a data usage agreement.

Specific to this study, subject IDs and code are available at https://github.com/tvanderwal/head_motion_2021.

http://fcon_1000.projects.nitrc.org/indi/scmi_healthy_brain_network/

https://github.com/tvanderwal/head_motion_2021

## Acknowledgements

The authors thank kinesiologist Jason Goode for helpful discussions. This work was supported by the BioTalent Canada Student Work Placement Program, the BC Children’s Hospital Research Institute Summer Studentship, and the BC Children’s Hospital Research Institute New Investigator Establishment Award.

## Notes

### Competing Interest Statement

The authors have declared no competing interest.

### Author Declarations

This study used publicly available data (Healthy Brain Network Biobank). All data collection for the HBN Biobank was approved by the Chesapeake Institutional Review Board (http://fcon_1000.projects.nitrc.org/indi/cmi_healthy_brain_network/).

